# Extraction and validation of patient housing and food insecurity status in a large electronic health records database using selective prediction and active learning

**DOI:** 10.1101/2022.12.06.22283140

**Authors:** Akshay Swaminathan, Wasan M Kumar, Ivan Lopez, Edward Tran, Ujwal Srivastava, William Wang, Max K Clary, Willemijn H van Deursen, Jonathan G Shaw, Olivier Gevaert, Jonathan H Chen

## Abstract

**Objective:** Information on patient social determinants of health is frequently recorded in unstructured clinical notes, making it inaccessible for researchers and policymakers. We aimed to extract and validate food and housing insecurity status on a large electronic health record-derived patient cohort by combining selective prediction and active learning.

**Materials and Methods:** Manually labeled charts selected via active learning were used to train L1-regularized logistic regression models to identify the presence of food insecurity (N=372, 42% event rate) and housing insecurity (N=559, 36% event rate) in clinical notes. In addition to validating predictions against labeled data, we further validated predictions on an additional unlabeled dataset through associative studies with demographic, clinical, and environmental variables with known associations with food and housing insecurity.

**Results:** The food insecurity model had AUC=0.83, sensitivity=0.90, PPV=0.90, and undetermined rate=0.59 (n=149); the housing insecurity model had AUC=0.81, sensitivity=0.50, PPV=1, and undetermined rate=0.65 (n=224). Out of 4,337 unlabeled patients, the 395 (9%) patients predicted to have food insecurity were more likely to be Hispanic/Latino (48% vs 24%, p<0.001) and have diabetes (34% vs 12%), hypertension (43% vs 11%), and heart disease (12% vs 0.7%) (p<0.001 for all).

**Discussion:** Selective prediction and active learning can facilitate efficient labeling of social determinants of health from unstructured EHR data to identify vulnerable populations and targets for healthcare system and policy intervention.

**Conclusion:** Machine learning can be used to extract high-fidelity information on patient food and housing insecurity status.

## Introduction

Social determinants of health (SDOH)—including access to housing, transportation, food, and access to healthcare—significantly influence health outcomes and alter the course of clinical care.^1^ SDOH factors have been linked to several health outcomes, including increased risk for chronic conditions, hospitalization, and mortality. Healthcare systems have sought to develop SDOH interventions but may struggle to identify patients with SDOH needs.^1,2^ To address this challenge, the National Academy of Medicine recommended that SDOH factors should be incorporated into Electronic Health Records (EHRs) to allow for improved research, policy, and quality improvement, spurring the development of a wide range of social risk screening tools.^3^

Despite the increasing use of social needs screens at point-of-care settings, most SDOH factors are captured as unstructured data, such as clinical notes, which can be difficult to analyze quantitatively. Traditional methods to identify SDOH needs include manual chart review, which is time-consuming. Advances in natural language processing (NLP) allow for the development of more efficient, automated and semi-automated approaches to unlock insights from unstructured clinical EHR notes. NLP models to study unstructured SDOH data may inform resource allocation and interventions by healthcare systems and can be used to improve risk stratification, clinical decision support algorithms, and screening tools.

Housing is one such SDOH factor that can significantly affect health, especially for patients with chronic conditions. At the individual level, housing insecurity reduces a person’s ability to care for conditions that require frequent monitoring or medication adherence. Given the well-researched link between housing and health, government and healthcare systems have begun to adopt housing interventions, which are shown to improve health outcomes and reduce costs.^4^ Housing status is frequently under-captured as structured data in EHR and clinical visits. In a study of a large EHR cohort of 1.2 million patients, approximately 3% of patients (35,000) had housing issues that were captured in unstructured data, compared to 0.19% of patients whose housing needs were in a structured format.^5^

Food insecurity is another SDOH factor impacting health, particularly through nutritional inadequacy, yet few studies have aimed to quantify the presence in EHR. For example, a recent systematic review of NLP extraction of SDOH variables identified only one paper that used machine learning (ML) methods to assess food insecurity, and this study did not use a naive test set and did not correct for negation or misspellings.^6,7^

Several chronic conditions are adversely affected by housing and food insecurity status, including coronary artery disease (CAD), hypertension (HTN), Type 2 Diabetes Mellitus (T2DM), hepatitis, and stroke.^8–12^ The relationship between diet and these chronic medical conditions is well-established. Adult patients who are food insecure, or have limited access to safe, adequately nutritious food, are more likely to have an increased risk for hypertension, with one study reporting a 21% increase in risk. Food insecurity is also associated with an increased prevalence of diabetes, as well as the co-occurrence of multiple chronic conditions.^4^ Food insecurity is associated with a higher intake of lower nutrition-quality food, thereby leading to higher rates of HTN and CVD. Patients who are housing insecure report poorer physical health outcomes, including hypertension and cardiovascular disease. One hypothesized mechanism is that patients who are housing insecure are less likely to access healthcare and preventive services, given their higher priority housing needs.^4^

Previous studies have used NLP models to assess SDOH status from clinical notes and have been able to abstract information on substance use, housing status, smoking status, alcohol use, social support, education, and financial needs.^7^ However, these studies face some important limitations. First, few studies have explored using ML and NLP to extract food insecurity status. Second, existing ML approaches to extract SDOH data do not adequately address variable model accuracy and the expensive process of data labeling. Third, few studies that use NLP to extract SDOH data apply models to unlabeled data and validate the resulting predictions.

This study aims to identify social risk factors (i.e. food insecurity and housing insecurity) in unstructured EHR data and using an NLP model and prospectively predict the presence of social risk in a patient’s EHR. Our approach addresses these limitations by using selective prediction, which gives prediction models to abstain from generating a prediction on uncertain charts, thus minimizing misclassifications; by selecting charts via active learning, which increases the efficiency of data collection by prioritizing charts that the model could most effectively learn from; by applying our model to housing insecurity and food insecurity, an understudied SDOH variable; and by validating model predictions using demographic (gender, race, ethnicity), geographic (zip codes), and clinical (diabetes, hypertension, heart disease, etc.) variables with known associations with food and housing insecurity. Our models can help researchers, providers, and policymakers identify patients with food and housing insecurity at scale.

## Methods

### Approach

First, manual abstraction of SDOH variables was performed by human abstractors (Figure 1a). Second, natural language processing (NLP) selective prediction models were developed using the data collected by manual abstraction. In contrast to typical ML models that generate a prediction for any input, selective prediction models may abstain from generating a prediction for specific data points.^13^ Third, we leveraged selective prediction modeling to generate predictions for data points that the model can predict with high accuracy and used active learning via uncertainty sampling to improve the efficiency of manual abstraction. Lastly, fully trained models were applied to unlabeled data and the resulting predictions were combined with manually labeled data to create an SDOH dataset used for downstream analysis.

**Figure 1:**
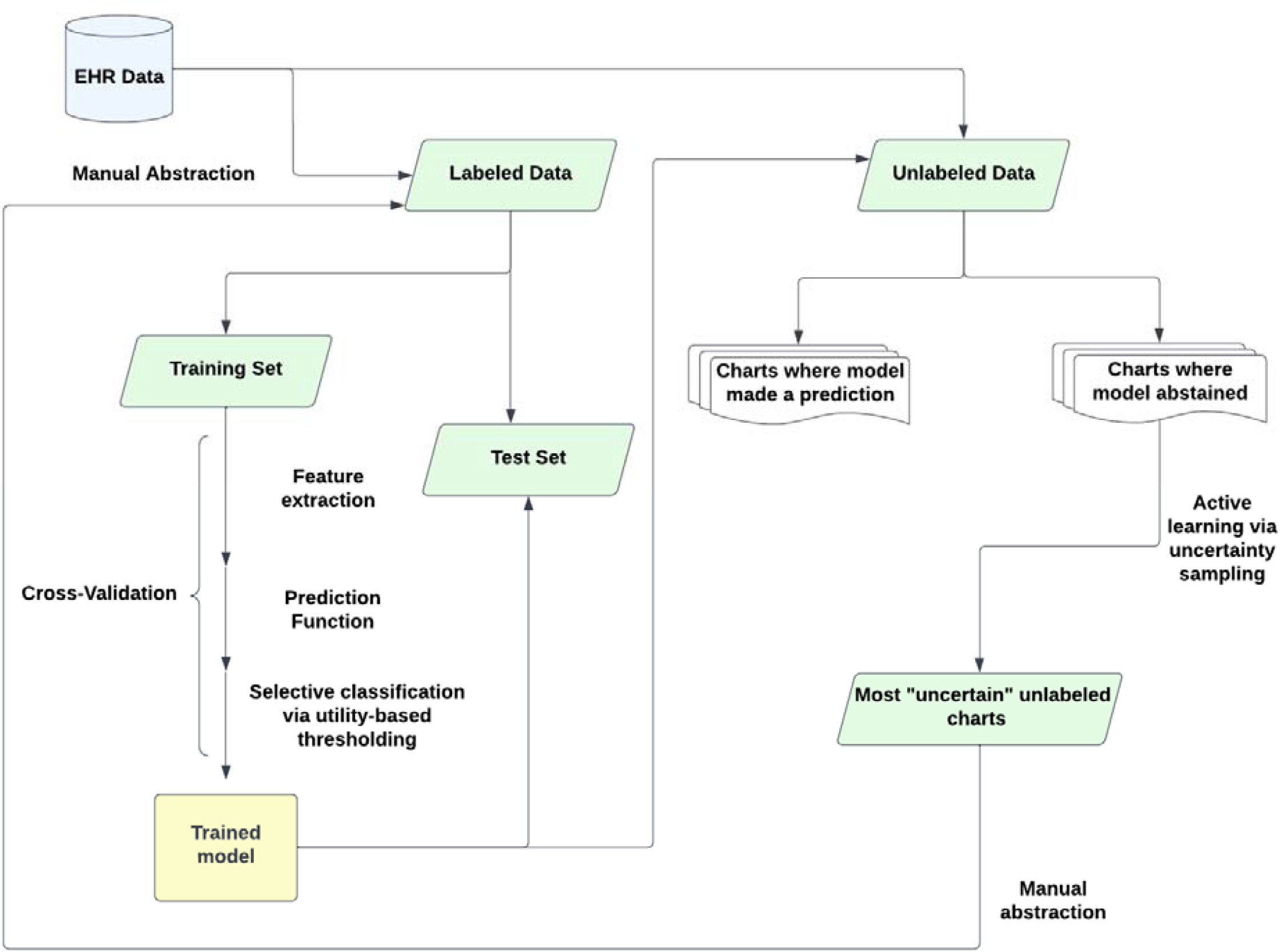
Diagram depicting the modeling workflow used to extract unstructured variables. EHR data is abstracted, then used to train and test a model that either predicts an outcome or abstains from a prediction. Charts for which a prediction is not generated are passed through an active learning framework via uncertainty sampling. These charts are then labeled and incorporated into the labeled dataset. This process was repeated twice for a total of three successively increasing labeled datasets.

**Figure 2a:**
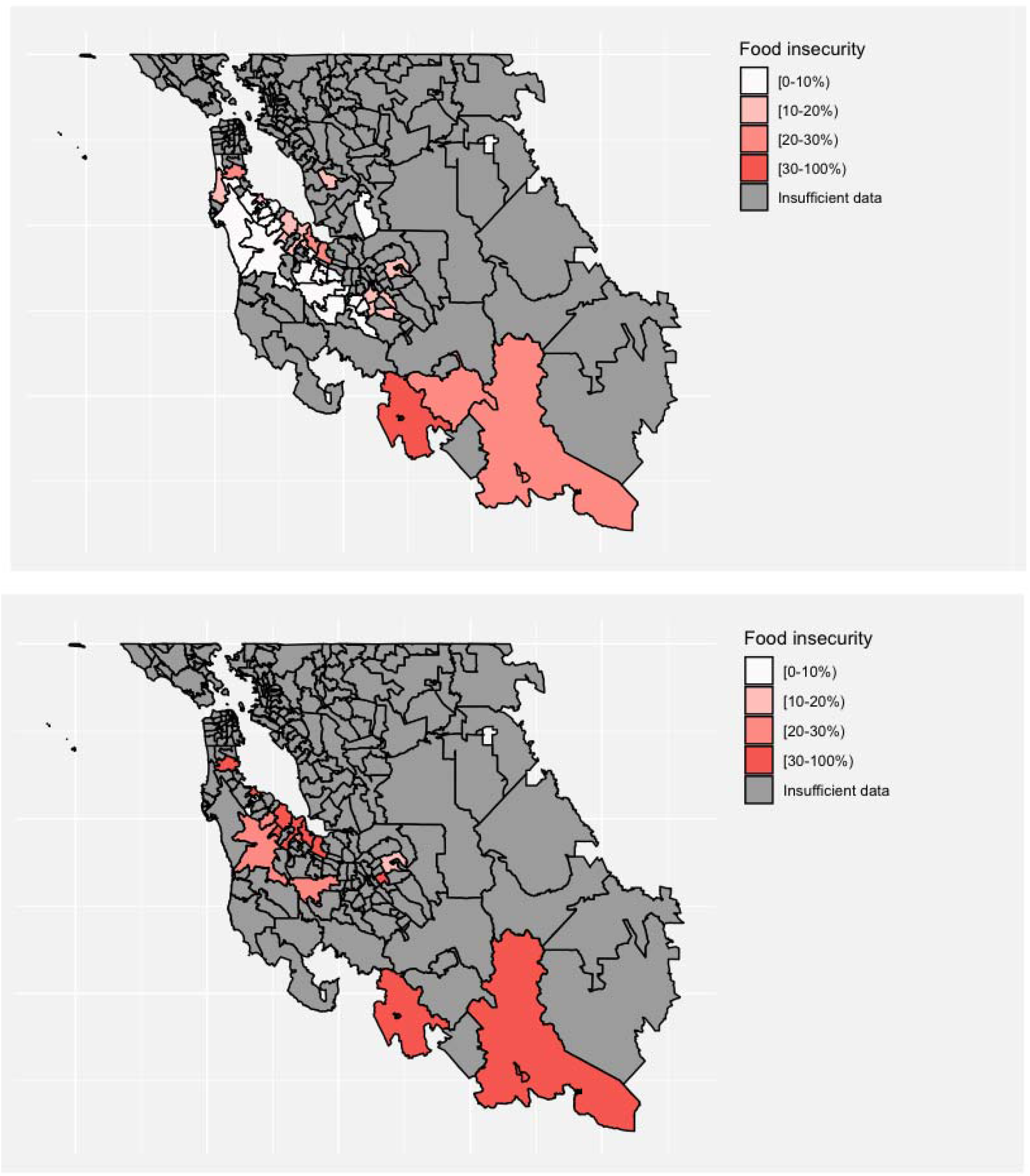
Map of predicted (top) and labeled (bottom) prevalence of food insecurity for Bay Area zip codes. Zip codes with at least 15 patients for the predicted cohort and at least 5 patients for the labeled cohort were included.

**Figure 2b:**
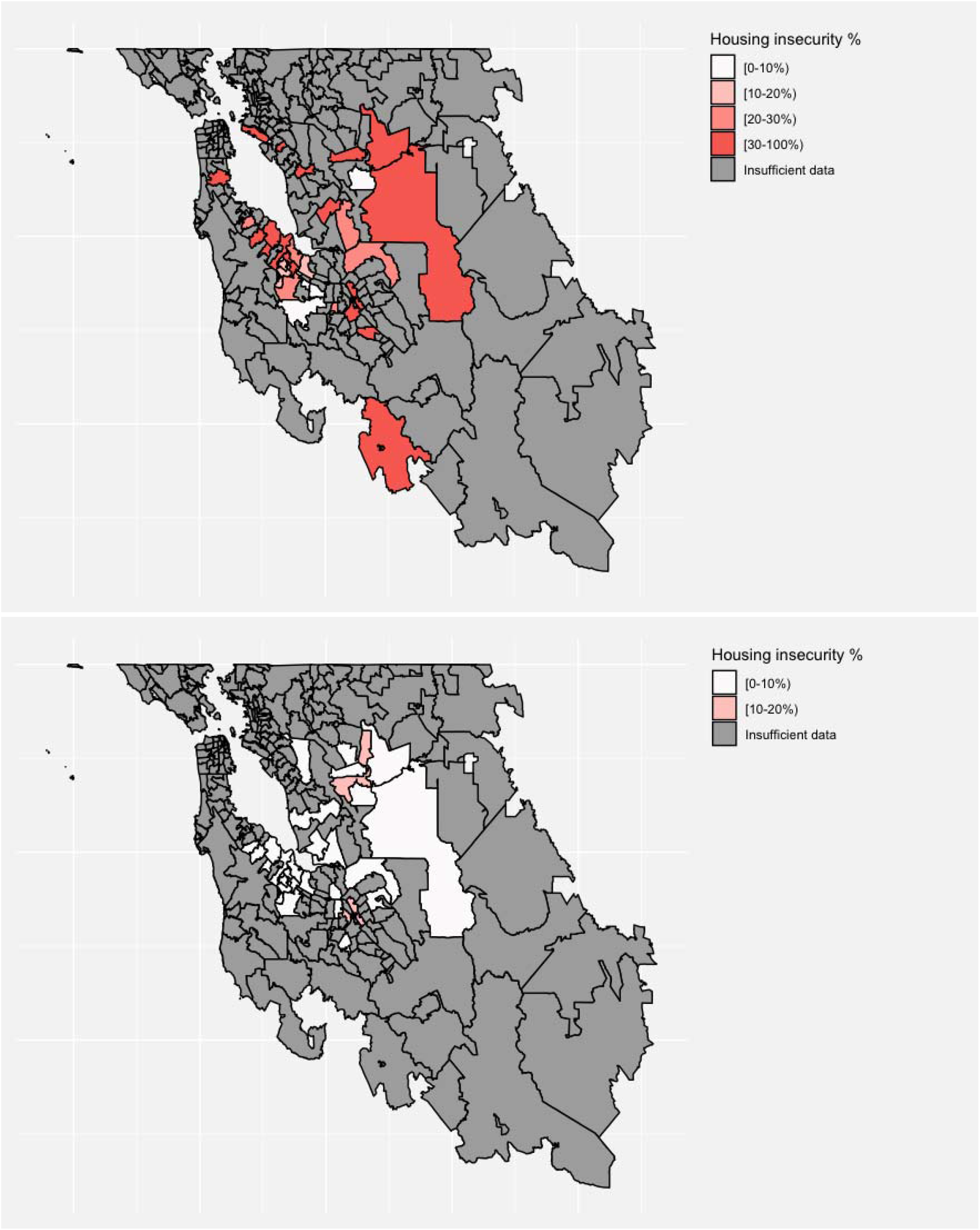
Map of predicted (top) and labeled (bottom) prevalence of housing insecurity for Bay Area zip codes. Zip codes with at least 15 patients for the predicted cohort and at least 5 patients for the labeled cohort were included.

### Data source

This study used a retrospective analysis of EHR data collected at Stanford Hospital, Stanford, CA, with data including any patient treated at Stanford Hospital from 1998-2022. Ethics approval was granted through Stanford University IRB (#65834).

### Study Population

#### Housing cohort

Patients with mention of the word “unhoused”, “shelter”, “homelessness” or “housing” in a clinical note between November and December 2022 were included. Clinical notes with these phrases were selected as they had a high likelihood of identifying individuals with housing needs. The remaining 1,705 patients were considered for subsequent analyses. Of these, 400 patients were selected for initial manual abstraction. Additional phrases described in the literature for housing insecurity, including “street” or “incarcerated”, led to a significant number of false positives due to improper settings (e.g. “supervise all play on streets/in driveways” or “incarcerated hernia”). We omitted these phrases to conserve model fidelity.

#### Food insecurity cohort

The food insecurity cohort was created by selecting patients that had a mention of the phrases “hunger”, “not enough food”, “food insec”, “food bank”, “food pantr”, “food stamp” in a clinical note between December 2010 and December 2022. The remaining 4,709 patients were considered for subsequent analyses. Of these, 200 patients were selected for initial manual abstraction.

### Outcomes

#### Food insecurity

Food insecurity was defined as any mention of limited access to nutritionally adequate and safe food, including patients who reported lack of food, consistent hunger, or inability to afford food; or patients receiving food stamps or other financial assistance for obtaining food (e.g. WIC, CalFresh program).^14,15^ This definition included individuals with both active food insecurity and a past history of food insecurity.

#### Housing insecurity

Housing insecurity was defined as any mention of inconsistent access to safe housing (e.g. homelessness, unstable housing). This included individuals with both active housing insecurity and a past history of housing insecurity.^4^

### Data abstraction

Of the patients that met the initial inclusion criteria, 200 and 400 patients were randomly selected for manual abstraction for food insecurity and housing insecurity, respectively. First, a chart review was performed to understand how food and housing insecurity was documented in the EHR. Next, a standardized abstraction data entry tool was used in order to refine search terms that were predictive of housing and food insecurity. The data entry tool had a “flag” feature that abstractors could use to indicate charts for which they were not able to make a determination. Lastly, all flagged charts were reviewed and reconciled.

### Statistical Analysis

#### Feature engineering and feature selection

For each variable, modeling was performed at the level of a patient chart. A single chart was randomly selected per patient. For simplicity, we henceforth refer to the unit of analysis as the chart. A randomly selected 60% of charts were used for model training, and the remaining 40% were used for model testing. Features were derived entirely from the free text of clinical notes. Using the text of these documents, TF-IDF (term-frequency-inverse-document-frequency) scores were calculated for unigrams, bigrams, and trigrams. Briefly, TF-IDF is a method used to quantify the relative frequency of an n-gram (an n-word phrase) in a corpus of documents. For example, the TF-IDF score for the unigram “insecurity” in a patient’s clinical report is calculated by dividing the frequency of the word “insecurity” in that patient’s clinical report (term frequency) by the frequency of the word “insecurity” across all clinical reports across all patients (inverse document frequency). L1-regularized logistic regression (Lasso) was used for feature selection and prediction. Hyperparameters (regularization for Lasso and sparsity for document term matrix) were tuned using 10-fold cross-validation. The metric for cross-validation was misclassification cost (described below). Selected features were examined for sensibility and clinical meaningfulness (SI Table 7).

#### Selective prediction

Selective prediction allows a model to abstain from making a prediction under certain circumstances.^13,16^ Consider a prediction function, f, that generates a continuous prediction given an input observation x. Let g represent the abstinence function — a function that is 1 if the model generates a prediction and 0 if the model abstains. Let h represent the selective prediction function, defined as

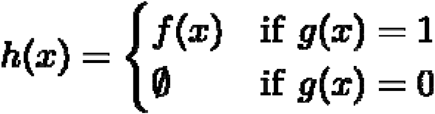

Where the empty set operator denotes the absence of a prediction. We henceforth describe this outcome as “undetermined”. In this study, f is the predicted probability of an L1-regularized logistic regression model and g is defined as follows:

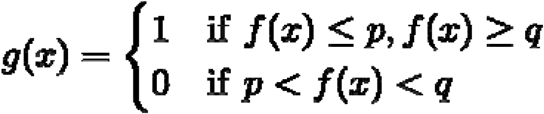

Where p and q represent distinct probability thresholds. Therefore, the resulting selective classifier, c, is defined as:

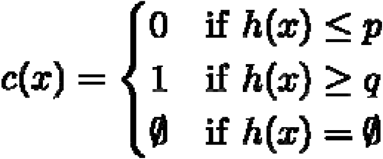

Following a previously described methodology for selective prediction via utility-based thresholding^17^, we select p and q to minimize the total cost of misclassification, defined as

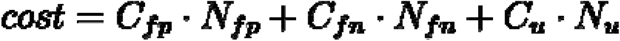

where C is cost, N is the number of observations, and the subscripts FP, FN, and U refer to false positives, false negatives, and undetermined, respectively. Here, “cost” refers not to monetary cost but rather real-world utilities. In the example of food insecurity, the cost of a false positive corresponds to the cost of incorrectly labeling a chart as having food insecurity, and the cost of a false negative corresponds to the cost of incorrectly labeling a chart as not having food insecurity. The former results in incorrectly including a patient in the final dataset, whereas the latter results in incorrectly excluding a patient from the final dataset. Following a previously-established regret-based approach, we set the relative costs of FP:FN:undetermined to be 13.5:8:1.^17^ This means that the model returning a false positive is 13.5 times more costly than abstaining. Similarly, a false negative result is 8 times more costly than the model abstaining.

#### Model evaluation

Area under the receiver operator curve (AUC), sensitivity, specificity, positive predictive value (PPV), and negative predictive value (NPV) were calculated, treating the abstracted labels as ground truth. For selective prediction models, we calculated undetermined rate, positive undetermined rate, and negative undetermined rate^17^. Positive undetermined rate is defined as the number of positive observations labeled as undetermined (undetermined positives or UP) over the total number of positive observations (TP + FN + UP). Similarly, the negative undetermined rate is defined as the number of negative observations (UN) labeled as undetermined over the total number of negative observations (TN + FP + UN). The undetermined rate is defined as the number of predictions a selective prediction model abstains from making (UN + UP), over the total number of possible predictions (UN + UP + FN + FP + TN + TP). A selective prediction model with a lower undetermined rate yields greater efficiency gains than a model with a larger undetermined rate and similar levels of accuracy.

After training, the final model was applied to all unlabeled data, yielding three types of predictions: “positive”, “negative” or “undetermined”. For charts labeled positive or negative, the model’s prediction was considered final. Charts labeled “undetermined” were those for which the model abstained from making a definitive prediction.

#### Active learning via uncertainty sampling

We used uncertainty sampling, an active learning technique, to improve the efficiency of manual abstraction.^18^ Active learning involves “actively” selecting data points for training such that the model learns faster compared to random selection of training data points.^19^ The idea is that if N training data points are required to reach a certain model performance using random sampling, fewer than N data points would be required to reach the same model performance using active learning. In uncertainty sampling, training data points that maximize a predefined measure of model uncertainty are prioritized for labeling. The motivation behind this approach is that the model can improve fastest by learning from data points about which it is currently uncertain. After initial model training, we selected the 100 data points whose predicted probabilities were between the two probability thresholds p and q (and therefore undetermined) and furthest away from either threshold. This corresponds to picking the 100 undetermined data points with the highest uncertainty score *u*, defined as

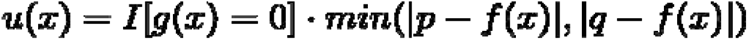

While it has been correctly pointed out that predicted probabilities are not a measure of model uncertainty, our goal was to help the model learn data points that fall in the undetermined region.^20^ Therefore, we defined u such that maximizing u would surface data points that fell squarely in the undetermined region. We performed two rounds of active learning following the initial round of model training.

#### Comparison of demographic and clinical characteristics

We compared relevant demographic and clinical characteristics between labeled and unlabeled datasets to ensure that the data used to develop the model (labeled) was similar to the data that the model was applied to (unlabeled). We also compared demographic and clinical characteristics between outcome groups (presence vs. absence of SDOH variable) in both the labeled and unlabeled (predicted) datasets to compare the actual (labeled) and predicted (unlabeled) relationships between the SDOH variable of interest and patient demographic and clinical characteristics. Demographic variables included patient gender, race, ethnicity, and need for an interpreter during a clinical encounter. Clinical variables were defined as the presence of at least one International Classification of Diseases (ICD) code for diabetes (insert codes), hypertension (insert codes), stroke (insert codes), hepatitis (insert codes), and heart disease (insert codes).

#### Geographic distribution of SDOH variables

To compare the actual and predicted geographic distribution of SDOH variables, we generated choropleth maps of the San Francisco Bay Area showing the prevalence of SDOH variables by patient zip code. To protect patient confidentiality, only zip codes with at least 5 patients were mapped. Shapefile data was obtained from Data SF.^21^

## Results

### Patient characteristics

All three cohorts of food insecure and housing insecure patients are similar for demographic and clinical characteristics (Table 1). The percentage of female patients in the training, test and unlabeled set was 50%, 50%, and 57%, respectively. The distribution across racial groups was similar in all three cohorts. The largest percentage of patients identified as Other (37% in the training set, 36% in the test set, 32% in the unlabeled set), followed by White (36%, 34%, 32%). The distribution across ethnicity was similar in all three cohorts. The largest percentage of patients identified as non-Hispanic (49% in the training set, 44% in the test set, 47% in the unlabeled set). Additionally, a similar percentage of patients needed an interpreter in all three cohorts (22%/23%/19% in train/test/unlabeled). The prevalence of diabetes (18%/17%/19% in train/test/unlabeled), hypertension (24%/27%/21%), heart disease (9.9%/3.4%/4.8%), and stroke (1.3%/2%/1.6%) were comparable in all three cohorts. The training and test cohorts had a similar prevalence of food insecurity (44% in train, 40% in test).

**Table 1:** Demographic and clinical characteristics of food and housing insecurity of training, testing, and unlabeled cohorts. Demographic variables include patient gender, race, ethnicity, and need for an interpreter during a clinical encounter. Clinical variables were defined as the presence of at least one International Classification of Diseases (ICD) code for diabetes (insert codes), hypertension (insert codes), stroke (insert codes), hepatitis (insert codes), and heart disease (insert codes).

|  | Food insecurity |  |  | Housing insecurity |  |  |
| --- | --- | --- | --- | --- | --- | --- |
| Characteristic | Test set, N = 149<sub>1</sub> | Training set, N = 223<sub>1</sub> | Unlabeled set, N = 4,337<sub>1</sub> | Test set, N = 224<sub>1</sub> | Training set, N = 335<sub>1</sub> | Unlabeled set, N = 1,166<sub>1</sub> |
| Outcome | 60 (40%) | 98 (44%) | - | 88 (39%) | 117 (35%) | - |
| **Gender** |  |  |  |  |  |  |
| Female | 75 (50%) | 112 (50%) | 2,459 (57%) | 124 (55%) | 174 (52%) | 701 (60%) |
| Male | 74 (50%) | 111 (50%) | 1,877 (43%) | 100 (45%) | 160 (48%) | 465 (40%) |
| Unknown | 0 (0%) | 0 (0%) | 1 (<0.1%) | 0 (0%) | 1 (0.3%) | 0 (0%) |
| **Race** |  |  |  |  |  |  |
| Asian | 11 (7.4%) | 24 (11%) | 384 (8.9%) | 39 (17%) | 47 (14%) | 212 (18%) |
| Black | 9 (6.0%) | 7 (3.1%) | 187 (4.3%) | 22 (9.8%) | 44 (13%) | 75 (6.4%) |
| Native American | 1 (0.7%) | 1 (0.4%) | 18 (0.4%) | 3 (1.3%) | 4 (1.2%) | 9 (0.8%) |
| Other | 54 (36%) | 82 (37%) | 1,397 (32%) | 47 (21%) | 76 (23%) | 278 (24%) |
| Pacific Islander | 2 (1.3%) | 2 (0.9%) | 68 (1.6%) | 2 (0.9%) | 7 (2.1%) | 16 (1.4%) |
| Unknown | 21 (14%) | 27 (12%) | 880 (20%) | 13 (5.8%) | 13 (3.9%) | 44 (3.8%) |
| White | 51 (34%) | 80 (36%) | 1,403 (32%) | 98 (44%) | 144 (43%) | 532 (46%) |
| Ethnicity |  |  |  |  |  |  |
| Hispanic/Latino | 63 (42%) | 82 (37%) | 1,451 (33%) | 39 (17%) | 56 (17%) | 224 (19%) |
| Non-Hispanic | 66 (44%) | 110 (49%) | 2,024 (47%) | 174 (78%) | 263 (79%) | 887 (76%) |
| Unknown | 20 (13%) | 31 (14%) | 862 (20%) | 11 (4.9%) | 16 (4.8%) | 55 (4.7%) |
| Interpreter needed |  |  |  |  |  |  |
| No | 114 (77%) | 174 (78%) | 3,509 (81%) | 211 (95%) | 305 (91%) | 1,081 (93%) |
| Yes | 35 (23%) | 48 (22%) | 805 (19%) | 10 (4.5%) | 30 (9.0%) | 85 (7.3%) |
| Recent BMI | 21 (17, 27) | 21 (17, 26) | 21 (17, 27) | 25 (22, 30) | 25 (21, 29) | 25 (22, 30) |
| Diabetes | 26 (17%) | 41 (18%) | 819 (19%) | 65 (29%) | 108 (32%) | 332 (28%) |
| Hypertension | 40 (27%) | 54 (24%) | 901 (21%) | 97 (43%) | 131 (39%) | 429 (37%) |
| Heart disease | 5 (3.4%) | 22 (9.9%) | 206 (4.8%) | 30 (13%) | 37 (11%) | 135 (12%) |
| Stroke | 3 (2.0%) | 3 (1.3%) | 71 (1.6%) | 14 (6.3%) | 23 (6.9%) | 43 (3.7%) |
| 7<br>n (%); Median (IQR) |  |  |  |  |  |  |

The percentage of female patients in the training, test and unlabeled set was 52%, 55%, and 60%, respectively. The distribution across racial groups was similar in all three cohorts. The largest proportion of patients identified as White (43% in the training set, 44% in the test set, 46% in the unlabeled set), followed by Other (23%, 21%, 24%). The distribution across ethnicity was similar in all three cohorts. The largest percentage of patients identified as non-Hispanic (79% in the training set, 78% in the test set, 76% in the unlabeled set). Additionally, a similar percentage of patients needed an interpreter in all three cohorts (9%, 4.5%, 7.3% in train/test/unlabeled). The prevalence of diabetes (32%/29%/28% in train/test/unlabeled), hypertension (39%/43%/37%), heart disease (11%/13%/12%), and stroke (6.9%/6.3%/3.7%) were comparable in all three cohorts. The training and test cohorts had a similar prevalence of food insecurity (35% in train, 39% in test).

### Effect of active learning on training and test set composition

The proportion of food insecure patients increased in the labeled datasets via the use of active learning (Table 2). In the initial labeled set (Round 1), 29% of patients had food insecurity, compared to 56% and 58% in the patients added via active learning in Rounds 2 and 3, respectively. Similarly, the use of active learning led to increases from Round 1 to Rounds 2 and 3 in the proportion of Hispanic/Latino patients (33% to 37% to 54%), non-English speaking patients (20% to 22% to 28%), and patients with chronic conditions.

**Table 2:** Comparison of demographic and clinical characteristics of food and housing insecurity of labeled cohort between initial training and test set (Round 1), patients added after one round of active learning (Round 2) and patients added after a second round of active learning (Round 3).

|  | Food insecurity |  |  | Housing insecurity |  |  |
| --- | --- | --- | --- | --- | --- | --- |
| Characteristic | Round 1, N = 190<sub>1</sub> | Round 2 additions, N = 99<sub>1</sub> | Round 3 additions, N = 83<sub>1</sub> | Round 1, N = 386<sub>1</sub> | Round 2 additions, N = 93<sub>1</sub> | Round 3 additions, N = 83<sub>1</sub> |
| Outcome | 55 (29%) | 55 (56%) | 48 (58%) | 110 (29%) | 42 (45%) | 53 (64%) |
| **Gender** |  |  |  |  |  |  |
| Female | 92 (48%) | 54 (55%) | 41 (49%) | 218 (57%) | 45 (48%) | 35 (42%) |
| Male | 98 (52%) | 45 (45%) | 42 (51%) | 165 (43%) | 48 (52%) | 47 (57%) |
| Unknown | 0 (0%) | 0 (0%) | 0 (0%) | 0 (0%) | 0 (0%) | 1 (1.2%) |
| **Race** |  |  |  |  |  |  |
| Asian | 19 (10%) | 11 (11%) | 5 (6.0%) | 69 (18%) | 12 (13%) | 5 (6.0%) |
| Black | 6 (3.2%) | 7 (7.1%) | 3 (3.6%) | 46 (12%) | 8 (8.6%) | 12 (14%) |
| Native American | 2 (1.1%) | 0 (0%) | 0 (0%) | 3 (0.8%) | 2 (2.2%) | 2 (2.4%) |
| Other | 59 (31%) | 31 (31%) | 46 (55%) | 76 (20%) | 20 (22%) | 27 (33%) |
| Pacific Islander | 3 (1.6%) | 1 (1.0%) | 0 (0%) | 7 (1.8%) | 0 (0%) | 2 (2.4%) |
| Unknown | 36 (19%) | 9 (9.1%) | 3 (3.6%) | 20 (5.2%) | 6 (6.5%) | 0 (0%) |
| White | 65 (34%) | 40 (40%) | 26 (31%) | 162 (42%) | 45 (48%) | 35 (42%) |
| Ethnicity |  |  |  |  |  |  |
| Hispanic/Latino | 63 (33%) | 37 (37%) | 45 (54%) | 55 (14%) | 18 (19%) | 22 (27%) |
| Non-Hispanic | 87 (46%) | 54 (55%) | 35 (42%) | 306 (80%) | 70 (75%) | 61 (73%) |
| Unknown | 40 (21%) | 8 (8.1%) | 3 (3.6%) | 22 (5.7%) | 5 (5.4%) | 0 (0%) |
| Interpreter needed |  |  |  |  |  |  |
| No | 151 (80%) | 77 (78%) | 60 (72%) | 349 (92%) | 87 (94%) | 80 (96%) |
| Yes | 38 (20%) | 22 (22%) | 23 (28%) | 31 (8.2%) | 6 (6.5%) | 3 (3.6%) |
| Recent BMI | 21 (17, 26) | 22 (18, 27) | 19 (15, 27) | 24 (21, 29) | 26 (22, 31) | 27 (23, 31) |
| Diabetes | 28 (15%) | 27 (27%) | 12 (14%) | 121 (32%) | 29 (31%) | 23 (28%) |
| Hypertension | 39 (21%) | 33 (33%) | 22 (27%) | 153 (40%) | 40 (43%) | 35 (42%) |
| Heart Disease | 10 (5.3%) | 12 (12%) | 5 (6.0%) | 44 (12%) | 11 (12%) | 12 (14%) |
| Stroke | 3 (1.6%) | 2 (2.0%) | 1 (1.2%) | 30 (7.9%) | 4 (4.3%) | 3 (3.6%) |
| n (%); Median (IQR) |  |  |  |  |  |  |

The proportion of housing insecure patients increased in the labeled datasets via the use of active learning (Table 2). In the initial training set (Round 1), 29% of patients had housing insecurity, compared to 45% and 64% in the added patients in Rounds 2 and 3, respectively. Similarly, the use of active learning led to increases from Round 1 to Rounds 2 and 3 in the proportion of Hispanic/Latino patients (14% to 19% to 27%), patients not requiring an interpreter (92% to 94% to 96%), and patients with chronic conditions.

### Model performance statistics

The selective prediction model for food insecurity had an AUC of 0.84, with high sensitivity (0.9), specificity (0.95), PPV (0.9) and NPV (0.95) (Table 3). The overall undetermined rate for predicting food insecurity was 59%, the positive undetermined rate was 67%, and the negative undetermined rate was 54%.

**Table 3:** Test set performance of classification models for food and housing insecurity. Note: positive undetermined rate = UP/(TP+FN+UP) and negative undetermined rate = UN/(TN+FP+UN), where UP (undetermined positives) is the number of positive observations labeled as undetermined; TP+FN+UP represents the total number of positive observations in the training dataset; UN (undetermined negatives) is the number of negative observations labeled as undetermined; and TN+FP+UN represents the total number of negative observations in the training dataset).

|  | Food Insecurity | Housing Insecurity |
| --- | --- | --- |
| Test set N | 149 | 224 |
| Event rate | 0.40 | 0.40 |
| AUC | 0.84 | 0.81 |
| Threshold 1 | 0.28 | 0.14 |
| Threshold 2 | 0.71 | 0.97 |
| Undetermined rate | 0.59 | 0.65 |
| Positive undetermined rate | 0.67 | 0.82 |
| Negative undetermined rate | 0.54 | 0.54 |
| Sensitivity | 0.90 | 0.50 |
| Specificity | 0.95 | 1 |
| PPV | 0.90 | 1 |
| NPV | 0.95 | 0.89 |

The selective prediction model for housing insecurity had an AUC of 0.81, with moderate sensitivity (0.5), high specificity (1), high PPV (1) and high NPV (0.89) (Table 3). The overall undetermined rate for predicting food insecurity was 65%, the positive undetermined rate was 82%, and the negative undetermined rate was 54%.

### Validation of predicted food insecurity status

The associations between food insecurity and demographic and clinical variables were similar in the unlabeled set compared to the labeled set. In the unlabeled set, patients with predicted food insecurity were more likely to be male (47% vs 41%, p=0.027), to be Hispanic/Latino (48% vs 24%, p<0.001), to need an interpreter (33% vs 11%, p<0.001), to have a higher BMI (23 vs 20, p<0.001), and to have diabetes (34% vs 12%), hypertension (43% vs 11%), heart disease (12% vs 0.7%), and stroke (3.3% vs 0.6%) (p<0.001 for all) (Table 4). This is similar to the labeled set, where patients with predicted food insecurity were more likely to be Hispanic/Latino (46% vs 34%, p<0.001), to need an interpreter (31% vs 16%, p<0.001), to have hypertension (37% vs 17%, p<0.001), and to have heart disease (11% vs 4.2%, p=0.008) (SI Table 2).

**Table 4:**
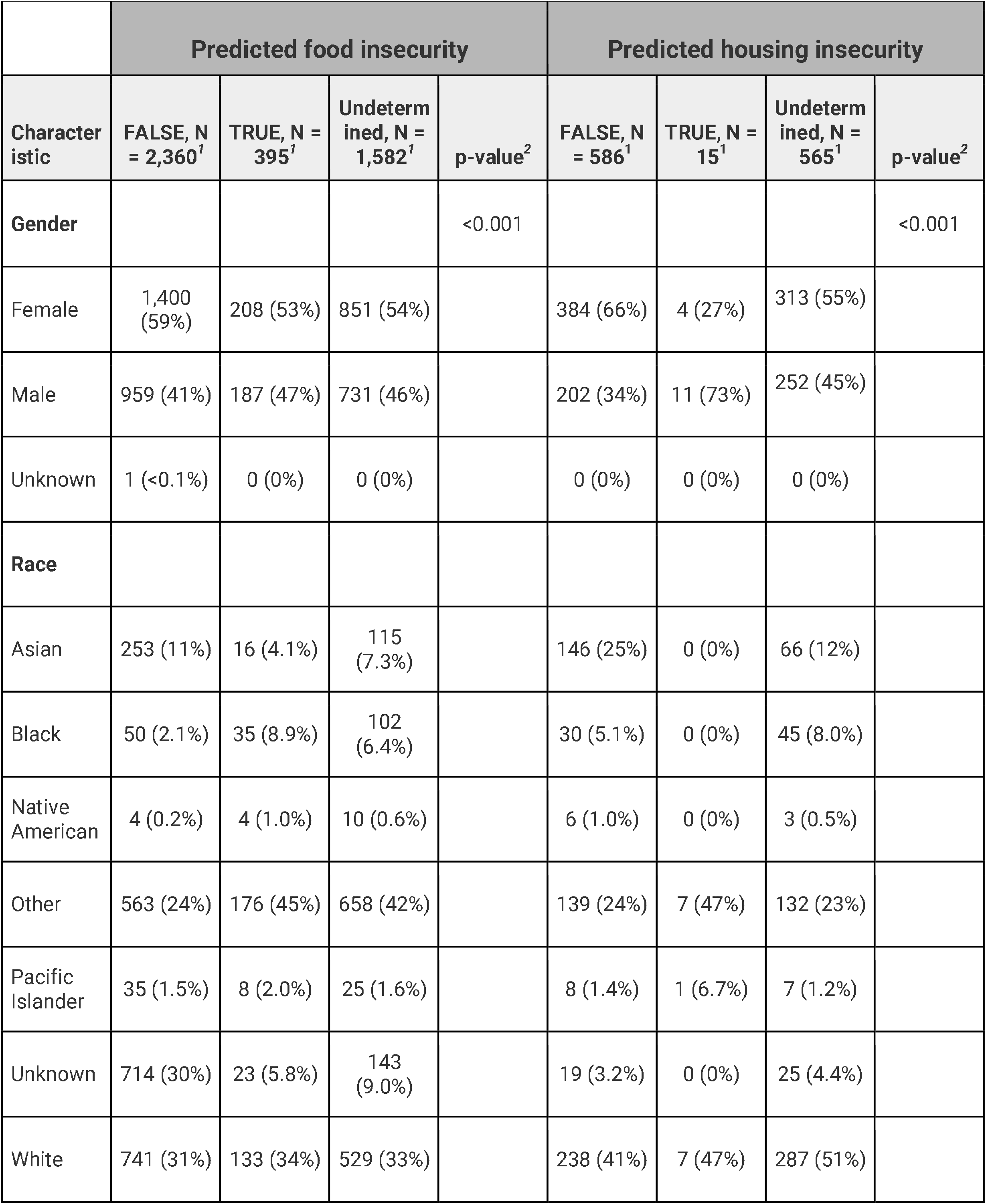
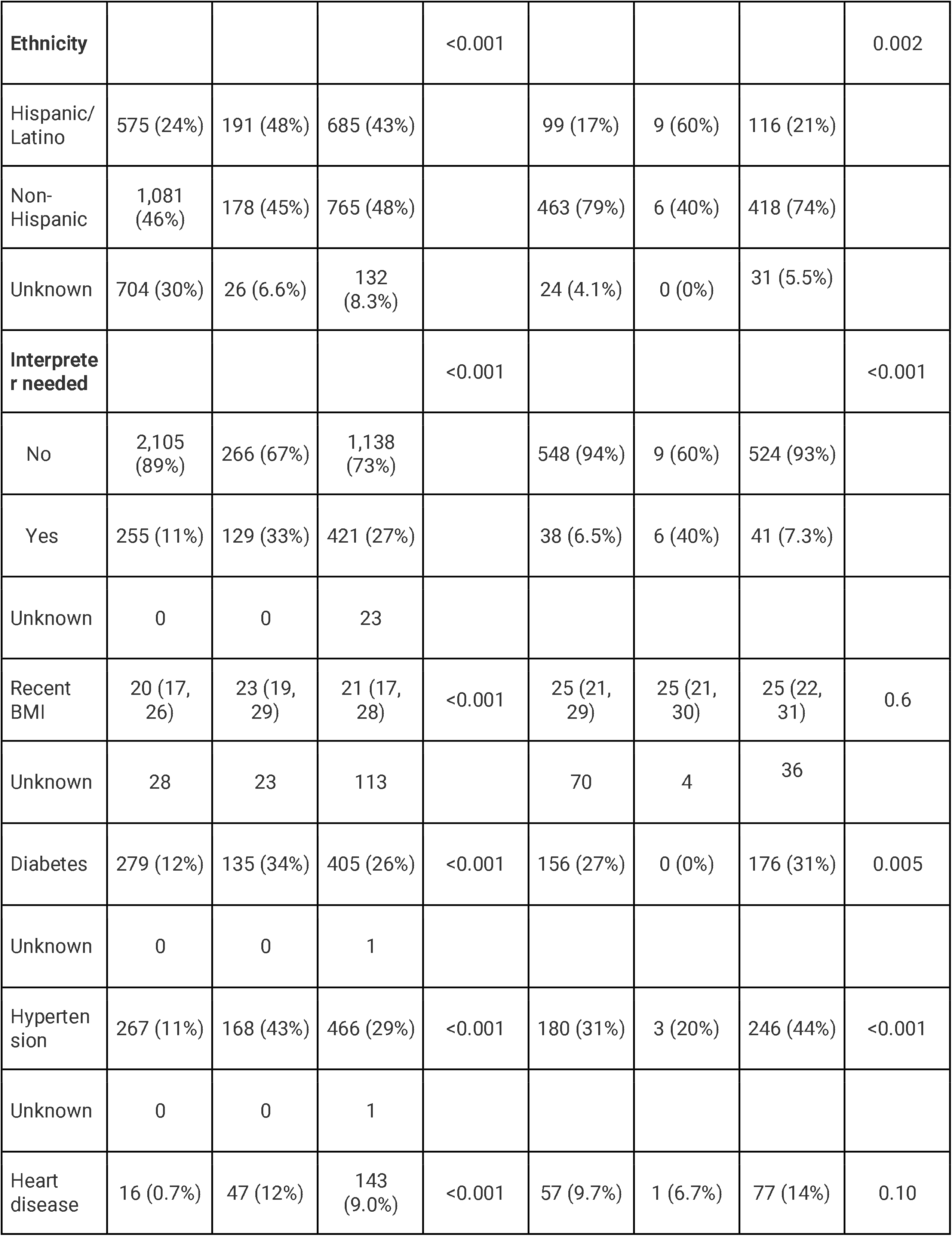
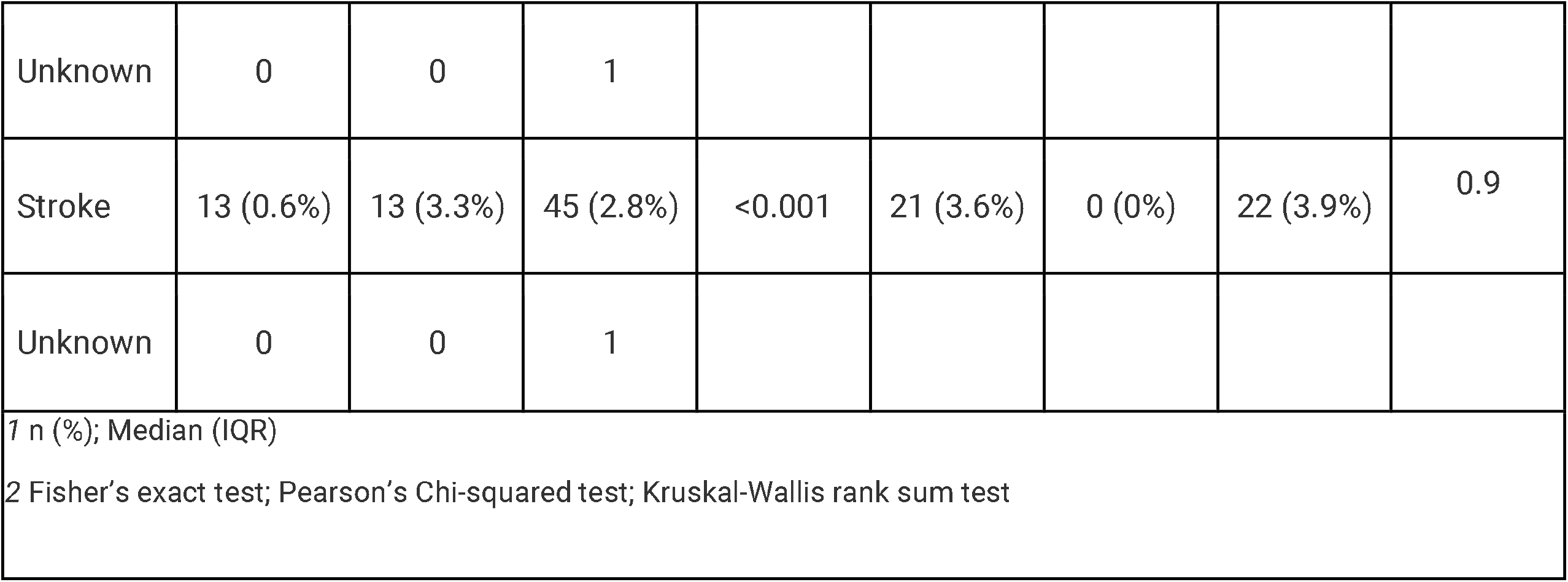
Comparison of demographic and clinical characteristics between patients with (TRUE) and without predicted food and housing insecurity in the unlabeled dataset. Patients for whom the model abstained from making a prediction are marked as “Undetermined”.

In the unlabeled set, patients with predicted housing insecurity were more likely to be male (73% vs 34%, p<0.001), to be Hispanic/Latino (60% vs 17%, p<0.001), to need an interpreter (40% vs 6.5%, p<0.001), and to not have diabetes (0% vs 27%, p=0.005) and hypertension (20% vs 31%, p<0.001) (Table 4). In the labeled set, patients with predicted housing insecurity were more likely to be male (59% vs 40%, p<0.001) and have higher BMI (26 vs 24, p=0.003) (SI Table 2).

Overall, the geographic distribution of food and housing insecurity was similar between the predicted dataset (unlabeled) and the labeled dataset. The zip codes with highest prevalence of predicted food insecurity were 95076 (48.8%), 94303 (25%), and 95020 (23.8%) (SI Table 3). In the labeled dataset, the zip codes with highest prevalence of predicted food insecurity were 95023 (60%), 95076 (60%), and 94303 (52.6%) (SI Table 4). Out of the 10 zip codes with the highest prevalence of food insecurity in the labeled set, 9 appeared in the top 10 zip codes with the highest predicted prevalence in the unlabeled set (SI Table 3,4).

The zip codes with highest prevalence of predicted housing insecurity were 95112 (16.7%), 95126 (16.7%), and 94588 (14.3%) (SI Table 5). In the labeled dataset, the zip codes with highest prevalence of predicted food insecurity were 94025 (62.5%), 95125 (55.6%), and 94080 (50.0%) (SI Table 6).

## Discussion

Our study demonstrates the feasibility of applying NLP models to unstructured clinical notes to identify patients with food and housing insecurity. We report three main findings. First, our selective prediction models had strong discriminative performance on a naive test set, indicating that they were able to accurately identify patients with food and housing insecurity. Second, the use of active learning via uncertainty sampling enriched the training datasets for data points that the model had trouble learning (instances of food and housing insecurity). Third, the predicted food insecurity status in large, unlabeled datasets yielded associations with demographic, geographic, and clinical variables that aligned both with associations in the labeled dataset, and with associations reported in the literature.

These findings suggest that this model is an accurate and efficient way to obtain SDOH information from an EMR. For example, we found that patients predicted to experience food insecurity were significantly more likely to have diabetes, hypertension, and stroke. This information could be utilized by physicians to inform an optimal treatment plan based on a patient’s specific social risks. Moreover, this data gives healthcare systems and quality improvement analysts a new opportunity to implement specific screenings and programs for the vulnerable populations identified by the model. In addition, little is known about the intersection of multiple SDOH and the impact on health, which this modeling approach could be used to better understand. While this study focused on food and housing insecurity, this model could be applied to numerous other SDOH, including substance use status, exposure to violence, disability status/access needs, transportation barriers, financial strain, among others.^22,23^ Our model makes previously difficult to quantify data on SDOH available to be addressed, which is an important first step towards achieving health equity.

Selective prediction is appropriate for use cases where not making a prediction is preferable to making an incorrect prediction. We previously showed how selective prediction can be applied for semi-automated cohort selection, where it is preferable to not make a prediction (and have a human manually abstract a chart, for example) than to incorrectly include or exclude a patient from a cohort.^17^ Here, we found that by not making a prediction on 40-50% of charts (undetermined rate), the model PPV was >95% on the remaining charts. The benefit of selective prediction via utility-based thresholding, as applied here, is that researchers can explicitly define their preferred trade-offs between making a prediction vs. abstaining.

Using active learning allowed us to identify high-yield charts for rapid model development. Active learning has been previously shown to lead to gains in labeling efficiency.^24^ Notably, both food insecurity and housing insecurity models identified groups of charts for active learning that tended to have higher event rates than the initial training sets. This suggests that the most “uncertain” charts were more likely to be events, and that using active learning helped the models more quickly learn these charts. We can approximate the impact of active learning on abstraction efficiency by comparing the number of events added through active learning with the number of charts required to abstract the same number of events through random sampling. For example, the food insecurity model added 55 + 48 = 103 events out of 182 total charts over the course of two rounds of active learning. Using the 29% event rate in the initial training set, it would have required abstracting 103 / 0.29 = ∼355 charts using random sampling to add the same number of events to the training data. This corresponds to a 51% reduction in the amount of abstraction required after applying active learning.

Our study is unique in its application of NLP models to unlabeled data and subsequent validation approaches. Although we did not abstract ground-truth labels for the unlabeled set, we showed that patients’ predicted SDOH status was correlated with demographic, geographic, and clinical variables in a manner consistent with both the labeled set and prior literature. For example, food insecurity is more prevalent in Black and Latino/Hispanic neighborhoods, which our model predicted.^25^ Similarly, food insecure adults in the U.S. are two to three times more likely to have diabetes than adults who are food-secure, consistent with our results 34% diabetes among food-insecure patients versus 12% diabetes among food-secure patients as predicted by the model.^26,27^ Lastly, patients experiencing housing insecurity were more likely to be from East Palo Alto and Salinas, which is consistent with literature showing the highest rates of homelessness in the San Francisco Bay Area are in Santa Clara County, where East Palo Alto and Salinas are located. These relationships suggest that our model-predicted SDOH variables recapitulate meaningful associations with demographic, clinical, and geographic variables.^21^ We note that the concordance of these associations was stronger for the food insecurity predictions than for housing insecurity prediction, suggesting that the housing insecurity variables may require further validation.

Future work could include accessing longitudinal data to understand chronic disease progression associated with SDOH factors and determining risks for chronic condition exacerbations. This would be particularly useful for clinicians in helping to determine the best treatment options for patients given their SDOH factors. For example, the model could be used to determine the likelihood of a person’s hypertension worsening given that they are food insecure. Additionally, it has been documented that several SDOH factors cluster and are associated with patient demographic characteristics.^30^ Future applications of the model could identify compound risk for chronic disease progression of multiple social determinants of health.

## Limitations

First, our training dataset utilizes a cohort of patients from a single institution for a one month period, which may have limited generalizability. Additional work should look to engage academic and non-academic healthcare institutions or publicly available data sources. Second, our model does not use a lexicon or ontology for identification, which may lead to a more comprehensive set of eligible patient charts. Third, manual abstraction focused on both a history of and active food and housing insecurity, which may have been complicated by uncertainties in clinical note entry (eg. an individual’s use of food stamp programs who does not consider themselves to be food insecure). Additionally, some chart data indicating food and housing insecurity might have been inaccessible because the formatting of the chart made it unreadable. Fourth, our model may be under-estimating the number of patients with food or housing insecurity given existing low-rates of capture for these variables in EMR.^28^ Since the cohort was limited to Stanford Healthcare, it is possible that the practices to obtain and report SDOH patient information within this system could vary from other healthcare practices. Fifth, we used a function of predicted probabilities from logistic regression for active learning, even though this metric is not strictly a measure of model uncertainty. Sixth, we did not conduct inter-rater reliability analyses for manually abstracted data, although abstractors were trained to follow a consistent definition and document unclear cases. Seventh, because the training dataset was enriched for events through active learning, the event rate in the unlabeled dataset might have been lower, resulting in miscalibrated predicted probabilities. Eighth, zip code data on patients with housing insecurity might not reflect their true zip code of residence as a result of unstable housing. Finally, the validation of chronic conditions was done using ICD codes which previously have been shown to have high specificity, but low sensitivity in determining true disease status.^29^

## Conclusion

Our study indicates that NLP is a feasible strategy to identify food and housing insecurity in patient EMR from clinical notes. Unlike previous studies, our model incorporates active learning and selective prediction, which allows for increased performance and training efficiency. Further, we used a unique tripartite validation strategy, which could be adopted to future studies using NLP to identify unstructured variables in the EMR. Our model showed strong test set accuracy and the predicted SDOH variables recapitulated known associations with demographic, geographic and clinical variables. We demonstrate this NLP approach to extract SDOH from patient EMR can successfully identify vulnerable populations and their specific risks, informing areas for healthcare system and policy intervention. Future studies should explore how NLP can be used to assess SDOH factors and risks for chronic condition progression and exacerbations, as well as the health effects of interventions to address SDOH needs.

## Data Availability

All data needed to evaluate the conclusions are present in the paper and in the Supplementary Materials.
The datasets generated analyzed during the current study are not publicly available due to patient privacy but are available from the corresponding author (JHC) on reasonable request.

## Supplementary Information

**SI Table 1:**
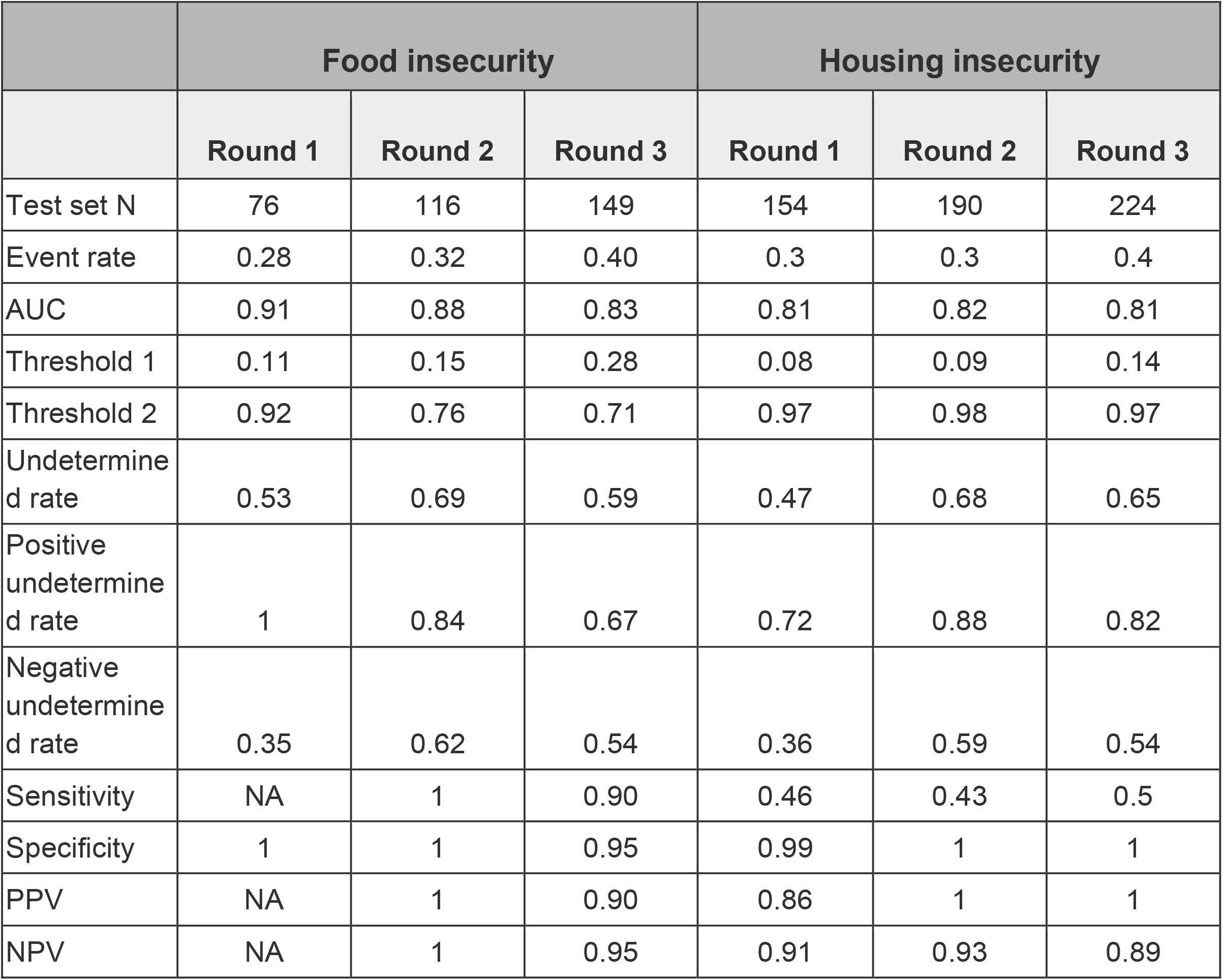
Performance metrics of test set for rounds 1, 2, 3 in predicting whether a patient is food insecure.

**SI Table 2:**
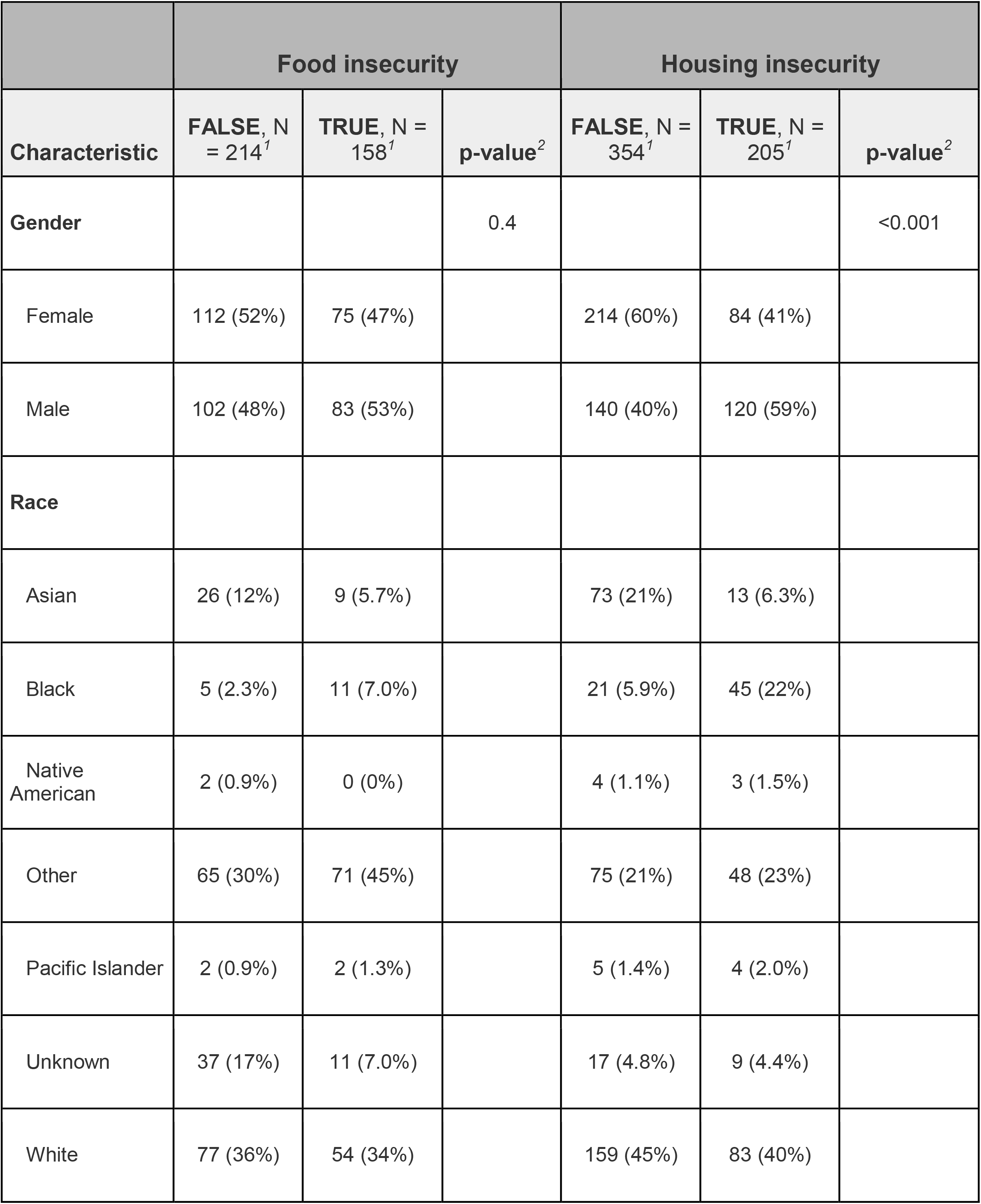
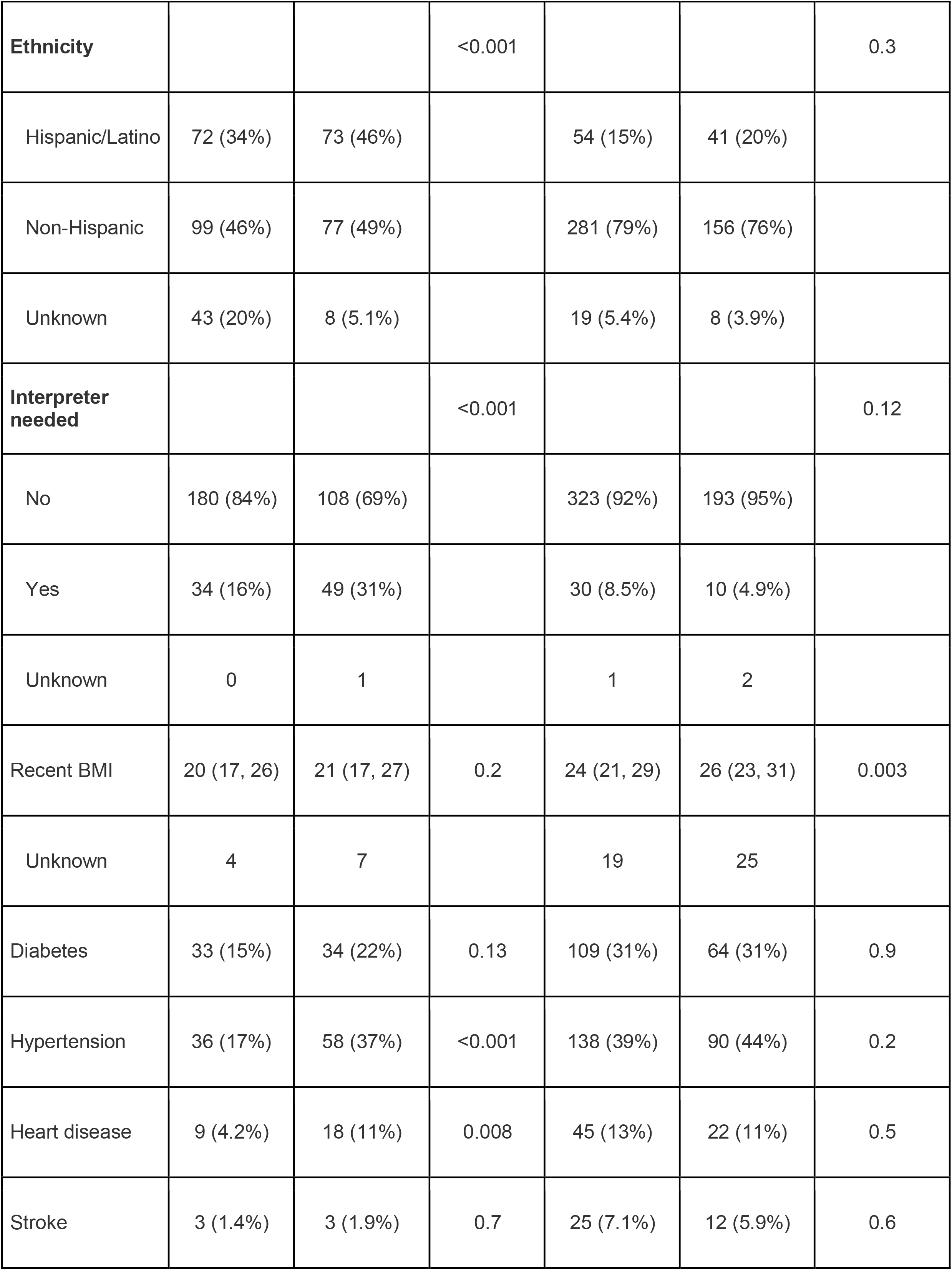
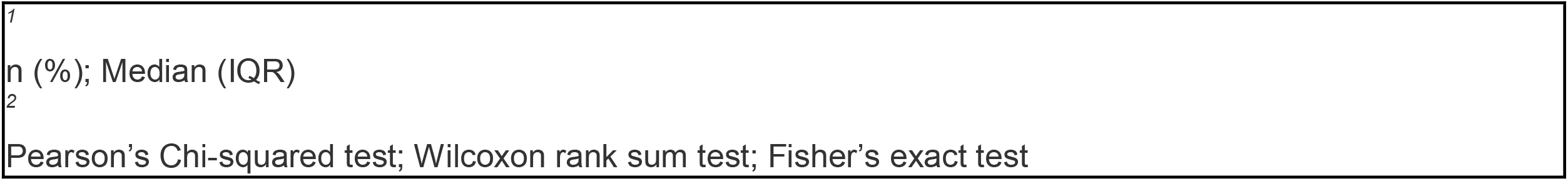
Comparison of demographic and clinical characteristics between patients with and without food insecurity in the labeled dataset (Round 3).

**SI Table 3:** This table contains the top 10 zip codes with the highest proportion of food insecurity produced by the unlabeled dataset. Zip codes are listed in descending order with the first zipcode as the highest proportion of food insecurity.

| Zip Code | City | Predicted Food Insecurity |
| --- | --- | --- |
| 95076 | Watsonville | 0.4884 |
| 94303 | East Palo Alto | 0.2500 |
| 95020 | Gilroy | 0.2381 |
| 95023 | Hollister | 0.2353 |
| 94043 | Mountain View | 0.2105 |
| 94080 | South San Francisco | 0.2051 |
| 94301 | Palo Alto | 0.1852 |
| 94063 | Redwood City | 0.1765 |
| 94061 | Redwood City | 0.1628 |
| 94025 | Menlo Park | 0.1605 |

**SI Table 4:** This table contains the top 10 zip codes with the highest proportion of food insecurity produced by the labeled dataset. Zip codes are listed in descending order with the first zipcode as the highest proportion of food insecurity.

| Zip Code | City Name | Food Insecurity |
| --- | --- | --- |
| 95023 | Hollister | 0.6000 |
| 95076 | Watsonville | 0.6000 |
| 94303 | East Palo Alto | 0.5263 |
| 94061 | Redwood City | 0.5000 |
| 94063 | Redwood City | 0.4667 |
| 94080 | South San Francisco | 0.4286 |
| 95122 | San Jose | 0.4286 |
| 94043 | Mountain View | 0.4000 |
| 94401 | San Mateo | 0.4000 |
| 94025 | Menlo Park | 0.3333 |

**SI Table 5:** This table contains the top 7 zip codes with the highest proportion of housing insecurity produced by the unlabeled dataset. Zip codes are listed in descending order with the first zipcode as the highest proportion of housing insecurity.

| Zip Code | City | Predicted housing insecurity |
| --- | --- | --- |
| 95112 | San Jose | 0.1667 |
| 95126 | San Jose | 0.1667 |
| 94588 | Pleasanton | 0.1429 |
| 94063 | Redwood City | 0.0833 |
| 94303 | East Palo Alto | 0.0476 |
| 94025 | Menlo Park | 0.0435 |
| 94550 | Livermore | 0.0323 |

**SI Table 6:** This table contains the top 10 zip codes with the highest proportion of housing insecurity produced by the labeled dataset. Zip codes are listed in descending order with the first zipcode as the highest proportion of housing insecurity.

| Zip Code | City | Housing insecurity |
| --- | --- | --- |
| 94025 | Menlo Park | 0.6250 |
| 95125 | San Jose | 0.5556 |
| 94080 | South San Francisco | 0.5000 |
| 94501 | Alameda | 0.5000 |
| 95076 | Watsonville | 0.4615 |
| 95112 | San Jose | 0.4444 |
| 94551 | Livermore | 0.4375 |
| 94568 | Dublin | 0.4286 |
| 94063 | Redwood City | 0.4000 |
| 95123 | San Jose | 0.4000 |

**SI Table 7:** Features and corresponding coefficients selected by the final food insecurity and housing insecurity L1-regularized logistic regression models.

| Food insecurity model |  | Housing insecurity model |  |
| --- | --- | --- | --- |
| Feature | Coefficient | Feature | Coefficient |
| no food | -1574.8 | total | -1201.8 |
| consistent | -971.51 | alcohol use | 1064.7 |
| %ile | -814.76 | glu | 941.02 |
| of care: | -760.87 | standard drinks | -936.09 |
| neurologic: | -752.68 | patient homeless? no | -920.87 |
| days of | -662.22 | level | 823.079 |
| tests | -580.08 | it was | 717.968 |
| frequency | -544.08 | includes | -708.41 |
| complete review | -537.19 | to keep | -705.48 |
| due | 390.458 | complete -point | -694.26 |
| screening: | -376.86 | assessment and | 638.515 |
| at least | 354.491 | and examined the | 562.628 |
| pertinent | -352.75 | call or | -558.61 |
| (last | -337.82 | in the | -550.83 |
| placement | 290.379 | spray | 546.744 |
| denies food insecurity. | -286.95 | sig | -528.4 |
| summary | 246.02 | type: | 527.991 |
| day | -235.03 | bolus | 526.076 |
| dinner: | -220.82 | anterior | 512.519 |
| sw | 220.392 | homelessness | 497.364 |
| procedures: | -183.91 | risk | -485.24 |
| highest | -183.33 | outpatient medications | -449.17 |
| to see | 148.585 | he has been | 415.976 |
| skin: | -147.86 | meth | 391.632 |
| need to | 143.76 | ). | -370.45 |
| social work | 136.763 | developed | -365.45 |
| old | -134.74 | / pulse | 325.035 |
| treatment | 126.243 | needs: | 322.691 |
| grossly | -124.55 | requesting | 296.942 |
| . | -112.46 | methamphetamine | 293.463 |
| this is | 99.2235 | worsening | -291.47 |
| chronic | -81.019 | note | 275.937 |
| otherwise | -79.18 | hospital | 259.638 |
| provided | 73.1336 | to participate in | 256.897 |
| patient is | 73.0803 | comprehensive | 225.496 |
| the patient | 71.8305 | mental status | 217.594 |
| file | -68.603 | homeless and | 208.87 |
| vitals: | -67.367 | diagnosis: | 208.071 |
| medication | 48.9694 | htn | -203.77 |
| /// | -33.479 | progress | 201.605 |
| air | -30.714 | spoke | 199.186 |
| complaint: | -27.281 | examined the | 194.095 |
| on file | -26.504 | please | -192.1 |
| distress | -23.648 | # | 186.685 |
| pt | 22.6803 | gap | 181.812 |
| continues | 12.7891 | mrn: | 180.846 |
| external | -10.209 | lower extremity | 180.797 |
| unable to | 5.0442 | resp | 174.937 |
| for evaluation | -1.3545 | was otherwise | -169.45 |
| use: | -0.9519 | manager | 162.855 |
| complete review of | 0 | but has | 153.104 |
|  |  | min: | 147.447 |
|  |  | intact. | 146.781 |
|  |  | number: | 146.21 |
|  |  | or more | -141.3 |
|  |  | reason: | 135.002 |
|  |  | shows | 133.45 |
|  |  | etoh | 131.161 |
|  |  | taken | 125.455 |
|  |  | vs: | -123.66 |
|  |  | abuse | 121.796 |
|  |  | july | 116.473 |
|  |  | questions | -114.88 |
|  |  | admit | 112.304 |
|  |  | thoughts, | -111.8 |
|  |  | stated | 110.494 |
|  |  | equipment | 110.317 |
|  |  | cooperative | 108.843 |
|  |  | he is | 108.153 |
|  |  | transplant | -105.21 |
|  |  | (pt | 102.092 |
|  |  | homeless | 98.8515 |
|  |  | he has a | 98.7985 |
|  |  | bed | 96.747 |
|  |  | going to | 90.4462 |
|  |  | °c ( . | 90.4029 |
|  |  | referring | 89.2087 |
|  |  | si/hi. | 86.0356 |
|  |  | independent | 84.5578 |
|  |  | progress note | 75.3714 |
|  |  | blood pressure | -74.756 |
|  |  | bleeding | 73.8246 |
|  |  | extra | -72.343 |
|  |  | use, | 71.1497 |
|  |  | please call | -70.267 |
|  |  | head | 67.8718 |
|  |  | acid | 61.4086 |
|  |  | attempted | 60.8398 |
|  |  | goals: | 58.7717 |
|  |  | ed | 52.5438 |
|  |  | mother, | -38.971 |
|  |  | minutes of | 36.2268 |
|  |  | assist | 28.467 |
|  |  | discharge | 27.4123 |
|  |  | performed during | 19.5393 |
|  |  | reported on | 19.2284 |
|  |  | """"a | 18.7031 |
|  |  | in a | -16.912 |
|  |  | diarrhea | 15.3017 |
|  |  | enjoys | -15.239 |
|  |  | sheltering** | -14.072 |
|  |  | unusual | -9.8525 |
|  |  | and affect | -6.3425 |
|  |  | sheltering** in | -4.6261 |
|  |  | you are | -3.9397 |
|  |  | medicine | 3.8918 |
|  |  | oriented, | -1.2078 |
|  |  | keep | -0.3927 |
|  |  | name | 0.1384 |
|  |  | performed during the | 0 |

## References

1. Integrating Social Care into the Delivery of Health Care: Moving Upstream to Improve the Nation’s Health.; 2019. doi:10.17226/25467

2. Ohanian A. The ROI of Addressing Social Determinants of Health. AJMC. Accessed December 5, 2022. https://www.ajmc.com/view/the-roi-of-addressing-social-determinants-of-health

3. Capturing Social and Behavioral Domains and Measures in Electronic Health Records: Phase 2.; 2014. doi:10.17226/18951

4. Taylor L. Housing And Health: An Overview Of The Literature | Health Affairs Brief. Accessed December 5, 2022. https://www.healthaffairs.org/do/10.1377/hpb20180313.396577/full/

5. Hatef E, Rouhizadeh M, Tia I, et al. Assessing the Availability of Data on Social and Behavioral Determinants in Structured and Unstructured Electronic Health Records: A Retrospective Analysis of a Multilevel Health Care System. JMIR Med Inform. 2019;7(3):e13802. doi:10.2196/13802

6. Stemerman R, Arguello J, Brice J, Krishnamurthy A, Houston M, Kitzmiller R. Identification of social determinants of health using multi-label classification of electronic health record clinical notes. JAMIA Open. 2021;4(3):ooaa069. doi:10.1093/jamiaopen/ooaa069

7. Patra BG, Sharma MM, Vekaria V, et al. Extracting social determinants of health from electronic health records using natural language processing: a systematic review. J Am Med Inform Assoc JAMIA. 2021;28(12):2716–2727. doi:10.1093/jamia/ocab170

8. Charkhchi P, Fazeli Dehkordy S, Carlos RC. Housing and Food Insecurity, Care Access, and Health Status Among the Chronically Ill: An Analysis of the Behavioral Risk Factor Surveillance System. J Gen Intern Med. 2018;33(5):644–650. doi:10.1007/s11606-017-4255-z

9. Bhat AC, Almeida DM, Fenelon A, Santos-Lozada AR. A longitudinal analysis of the relationship between housing insecurity and physical health among midlife and aging adults in the United States. SSM - Popul Health. 2022;18:101128. doi:10.1016/j.ssmph.2022.101128

10. Seligman HK, Laraia BA, Kushel MB. Food Insecurity Is Associated with Chronic Disease among Low-Income NHANES Participants. J Nutr. 2010;140(2):304–310. doi:10.3945/jn.109.112573

11. Leung CW, Kullgren JT, Malani PN, et al. Food insecurity is associated with multiple chronic conditions and physical health status among older US adults. Prev Med Rep. 2020;20:101211. doi:10.1016/j.pmedr.2020.101211

12. Yadav RS, Chaudhary D, Avula V, et al. Social Determinants of Stroke Hospitalization and Mortality in United States’ Counties. J Clin Med. 2022;11(14):4101. doi:10.3390/jcm11144101

13. Xin J, Tang R, Yu Y, Lin J. The Art of Abstention: Selective Prediction and Error Regularization for Natural Language Processing. In: Proceedings of the 59th Annual Meeting of the Association for Computational Linguistics and the 11th International Joint Conference on Natural Language Processing (Volume 1: Long Papers). Association for Computational Linguistics; 2021:1040–1051. doi:10.18653/v1/2021.acl-long.84

14. USDA ERS - Measurement. Accessed December 5, 2022. https://www.ers.usda.gov/topics/food-nutrition-assistance/food-security-in-the-u-s/measurement/

15. California Department of Social Services. CalFresh. Accessed December 5, 2022. http://www.cdss.ca.gov/inforesources/calfresh

16. Varshney N, Mishra S, Baral C. Towards Improving Selective Prediction Ability of NLP Systems. In: Proceedings of the 7th Workshop on Representation Learning for NLP. Association for Computational Linguistics; 2022:221–226. doi:10.18653/v1/2022.repl4nlp-1.23

17. Swaminathan A, Lopez I, Wang W, et al. Selective Prediction for Extracting Unstructured Clinical Data. Health Informatics; 2022. doi:10.1101/2022.11.15.22282368

18. Lewis DD, Gale WA. A Sequential Algorithm for Training Text Classifiers. Published online July 24, 1994. Accessed December 5, 2022. http://arxiv.org/abs/cmp-lg/9407020

19. Prince M. Does Active Learning Work? A Review of the Research. J Eng Educ. 2004;93(3):223–231. doi:10.1002/j.2168-9830.2004.tb00809.x

20. Gal Y, Ghahramani Z. Dropout as a Bayesian Approximation: Representing Model Uncertainty in Deep Learning. In: Proceedings of The 33rd International Conference on Machine Learning. PMLR; 2016:1050–1059. Accessed December 5, 2022. https://proceedings.mlr.press/v48/gal16.html

21. Bay Area ZIP Codes | DataSF | City and County of San Francisco. San Francisco Data. Accessed December 5, 2022. https://data.sfgov.org/Geographic-Locations-and-Boundaries/Bay-Area-ZIP-Codes/u5j3-svi6

22. Weir RC, Proser M, Jester M, Li V, Hood-Ronick CM, Gurewich D. Collecting Social Determinants of Health Data in the Clinical Setting: Findings from National PRAPARE Implementation. J Health Care Poor Underserved. 2020;31(2):1018–1035. doi:10.1353/hpu.2020.0075

23. Browse Objectives - Healthy People 2030 | health.gov. Accessed December 5, 2022. https://health.gov/healthypeople/objectives-and-data/browse-objectives

24. Yang Y, Loog M. A Benchmark and Comparison of Active Learning for Logistic Regression. Pattern Recognit. 2018;83:401–415. doi:10.1016/j.patcog.2018.06.004

25. New Poverty and Food Insecurity Data Illustrate Persistent Racial Inequities. Center for American Progress. Accessed December 5, 2022. https://www.americanprogress.org/article/new-poverty-food-insecurity-data-illustrate-persistent-racial-inequities/

26. Fitzgerald N, Hromi-Fiedler A, Segura-Pérez S, Pérez-Escamilla R. Food Insecurity is Related to Increased Risk of Type 2 Diabetes Among Latinas. Ethn Dis. 2011;21(3):328–334. Accessed December 5, 2022. https://www.ncbi.nlm.nih.gov/pmc/articles/PMC4048712/

27. Seligman HK, Bindman AB, Vittinghoff E, Kanaya AM, Kushel MB. Food insecurity is associated with diabetes mellitus: results from the National Health Examination and Nutrition Examination Survey (NHANES) 1999-2002. J Gen Intern Med. 2007;22(7):1018–1023. doi:10.1007/s11606-007-0192-6

28. Cook LA, Sachs J, Weiskopf NG. The quality of social determinants data in the electronic health record: a systematic review. J Am Med Inform Assoc JAMIA. 2021;29(1):187–196. doi:10.1093/jamia/ocab199

29. Goldstein N, Kahal D, Testa K, Gracely E, Burstyn I. Data Quality in Electronic Health Record Research: An Approach for Validation and Quantitative Bias Analysis for Imperfectly Ascertained Health Outcomes Via Diagnostic Codes. Harv Data Sci Rev. 2022;4(2). doi:10.1162/99608f92.cbe67e91

30. Schiltz NK, Chagin K, Sehgal AR. Clustering of Social Determinants of Health Among Patients. J Prim Care Community Health. 2022;13:21501319221113544. doi:10.1177/21501319221113543

